# Knowledge, Attitude, Perceptions and Practice towards COVID-19: A systematic review and Meta-analysis

**DOI:** 10.1101/2020.06.24.20138891

**Authors:** Akshaya Srikanth Bhagavathula, Wafa Ali Aldhaleei, Jamal Rahmani, Jagdish Khubchandani

**Author notes:** **Corresponding author** Dr. Akshaya Srikanth Bhagavathula, PharmD, PhD student, Institute of Public Health, College of Medicine and Health Sciences, UAE University, Al Ain, UAE.

## Abstract

**Background:** Several studies among various population groups have been conducted to investigate the level of knowledge, attitudes, perceptions, and risk reduction practices (KAP) related to COVID-19. A comprehensive review on this topic is important to highlight the areas for improvement and interventions to prevent COVID-19. Thus, the purpose of this study was to summarize the level of KAP about COVID-19 via a systematic review

**Methods:** A systematic literature search was performed using a combination of selected keywords in four scientific databases to identify relevant literature published from January 1 to May 31, 2020. Nineteen articles were included in the systematic review, and sixteen studies in the meta-analysis. The data was analyzed using a random-effects model due to the heterogeneity between the studies.

**Results:** Lack of COVID-19-related knowledge, positive perceptions, and preventive practices were detected and seems widespread. In particular, 56.6% (95%CI: 45.9-67%) of the health care workers (HCWs) and medical students had poor knowledge about COVID-19 and only 46% (95%CI: 15-77) of the total study sample had positive perceptions towards COVID-19. Besides, 81.7% of the sample prioritized practicing hand hygiene to prevent COVID-19, but wearing a face mask to prevent COVID-19 transmission was suboptimal (73.4%). Finally, around eighty percent of the subjects had good knowledge about COVID-19 symptoms (79%) and its transmission (82%) and reported that they avoided crowded places to prevent getting COVID-19 (89%).

**Conclusion:** Evidence-based practices on risk communication and raising awareness should be planned by local governments in collaboration with healthcare organizations. Specifically, educational initiatives for HCWs to prioritize wearing a face mask and practicing hand hygiene should be considered a priority.

## Introduction

The novel coronavirus (COVID-19) pandemic is a global health concern approaching 10 million cases by the end of June 2020 [1]. COVID-19 infection is highly transmissible causing profound social, economic, and political upheaval worldwide. So far, no antiviral drugs or vaccines are available explicitly to prevent or cure COVID-19 infection. Absent such measures, applying preventive strategies to control the infection are of paramount importance. Since the origin of COVID-19 in China, several measures were deployed by governments and public health organizations worldwide to raise awareness, improve knowledge, and to strengthen the preventive measures to control COVID-19 transmission [2-4]. However, lack of knowledge about COVID-19 transmission, inadequate understanding of the population at risk, and not paying attention to preventive measures are still widespread among regions and populations [5-8]. As a result, COVID-19 infections continue to spread and cause profound morbidity and mortality around the world. Several studies among various population groups across regions have been conducted to investigate the level of knowledge, attitudes, perceptions, and risk reduction practices as it relates to the prevention of COVID-19 transmission (KAP) [9-27]. Despite these studies, in a comprehensive review of literature, we could not find a systematic review summarizing global KAP on COVID-19 prevention practices. In the absence of such an evidence synthesis review, gaps in KAP and COVID-19 prevention practices around the world cannot be identified. In addition, such reviews are critical to provide empirical evidence on interventions and strategies that can be implemented for COVID-19 prevention among various populations. Thus, the purpose of this study was to conduct a systematic review and meta-analysis to summarize KAP on COVID-19 and to highlight areas for required interventions and evidence-based practices to prevent COVID-19.

## Methods

### Strategy

A systematic review and meta-analysis on KAP about COVID-19 following the Preferred Reporting Items for Systematic Review and Meta-Analysis (PRISMA) guidelines was conducted [28]. Cross-sectional observational studies investigating the knowledge, attitude, perceptions, and practice about COVID-19 and published from January – May 31, 2020, were included in our analysis.

### Literature Search

A literature search was conducted using MeSH keywords in four databases of peer-reviewed publications (PubMed, Scopus, Embase, and Google scholar). The following keywords were used: knowledge* OR attitude* OR perception* OR belief* OR practice*, AND cross-sectional studies*, AND questionnaire*, AND surveys*, AND observational* AND coronavirus* OR coronavirus infections*, OR novel coronavirus* OR covid-19*, OR severe acute respiratory syndrome* OR coronavirus disease* AND physicians* OR doctors* OR primary care* OR dentists* OR dental* OR nurses* OR nursing* OR community health workers* OR public health nursing* OR health professionals* OR public health* OR pharmacy* OR students* OR *general public OR population* OR community*. The field was limited to “title/abstract,” and the publication type was limited to “journal article” and “pre-prints” published in the English language. Letter to editors, case reports, study protocols, reviews, and interventional studies were not included in this study. The reference list of articles found in our search were also reviewed to identify other articles. We limited our review to the studies that used a structured questionnaire administered among different population assessing the following:

- Knowledge: COVID-19 symptoms
- Knowledge: COVID-19 transmission
- Attitude about COVID-19 isolation
- Avoiding crowded places to prevent the spread of COVID-19
- Perceptions about COVID-19
- Wearing facemask for COVID-19 protection
- Practicing hand hygiene

No published or in-progress systematic review on this topic was recognized in the Cochrane Library and PROSPERO before this review. The protocol for this systematic review and meta-analysis has been registered in PROSPERO 2020 (CRD42020188371) [29].

### Selection of Studies

Based on the aforementioned inclusion and exclusion criteria, articles using keywords related to the knowledge, attitude, perception, and practice about COVID-19 were selected. The study authors independently screened the titles and abstracts to identify eligible studies. Only full-text papers available in the English language were included. Small changes in the wording were also overlooked to understand their exact functional meaning. The authors excluded duplicates, articles not meeting inclusion/exclusion criteria, and studies in which the data were inadequately reported [Figure 1].

**Figure 1:**
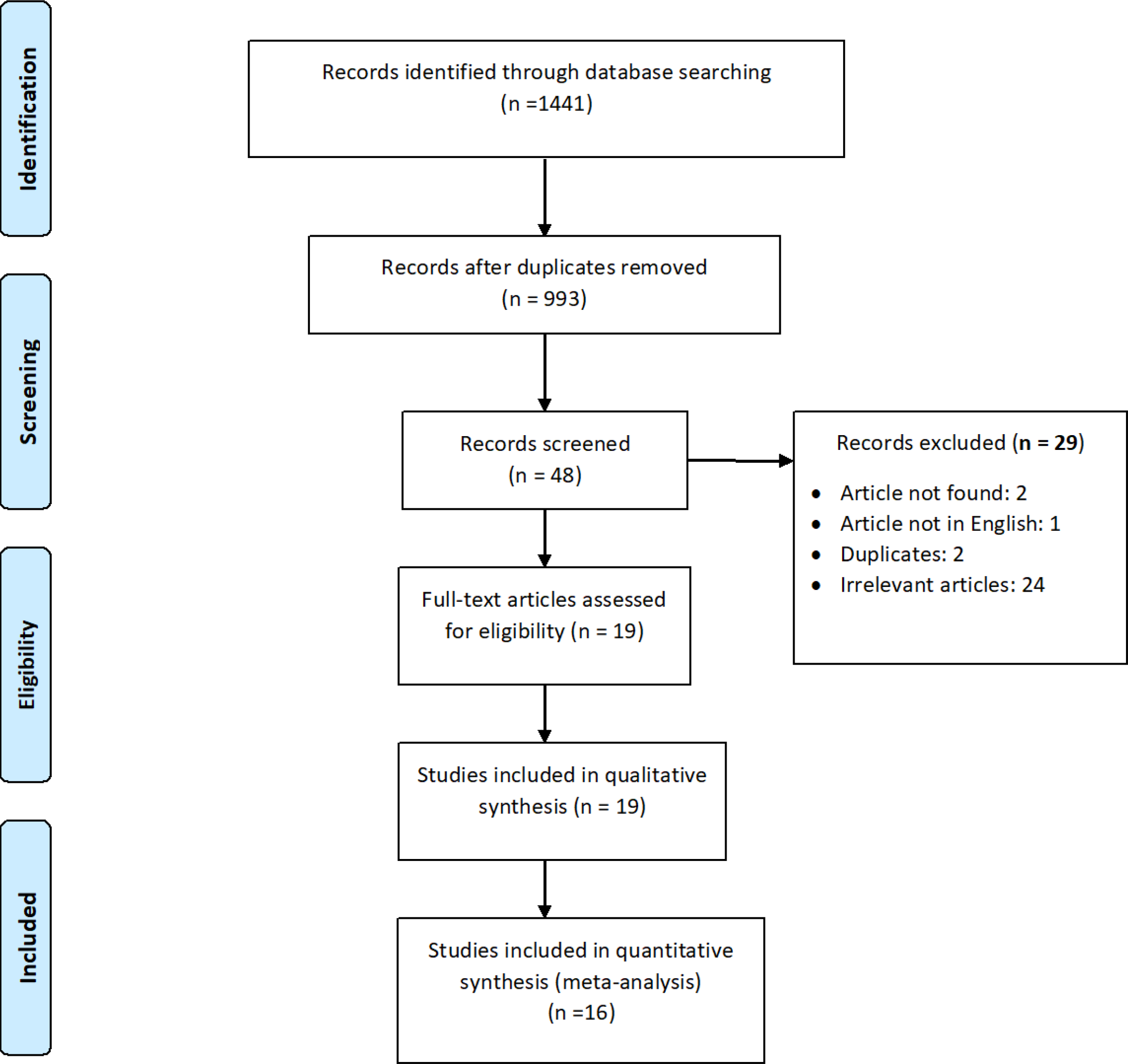
Flow of information through different phases of the systematic review.

### Data Extraction

The data for this review we extracted the following information such as author names, study design, study location, sampling, methods of administration of the questionnaire, and key results; these data were obtained from the selected articles and recorded in an Excel sheet. The data reported in or calculated from the included studies were used for analysis. Discrepancies related to the inclusion of particular studies were resolved by discussion among authors to reach consensus.

### Quality assessment

Methodological quality and risk of bias of each included study were evaluated using the Joanna Briggs Institute’s checklist for the critical appraisal (i.e., a nine-item checklist to evaluate that the sample is representative of the target population) [30]. Questions included the following: were the study participants recruited appropriately? was the sample size adequate? were the study subjects and settings described precisely? was data analysis used to identify the sample? were objectives and standard criteria used to measure the condition? and were important confounders identified or considered? The methodological quality of the studies was also assessed using the Strengthening the Reporting of Observational Studies in Epidemiology (STROBE) scale [31]. Egger and Begg tests and graphs representing funnel plots evaluated publication bias.

### Statistical analysis

Meta-analysis was performed using STATA version 16 software. The heterogeneity of the studies was evaluated using Cochrane’s Q-test and *I*^*2*^statistics. The random-effects model was used to combine the studies showing heterogeneity of Cochrane Q *p*<0.10 and *I*^*2*^ >50%. A forest plot was used to demonstrate the selected studies in terms of estimates with a 95% confidence interval (CI). The differences in knowledge, attitude, perception, and practice across various study groups were assessed using subgroup analysis. Sensitivity analysis was also performed by stratified the studies into high quality (over 75% of the STROBE checklist) and low quality (under 75% of the STROBE checklist).

## Results

A total of 1441 studies were found in the initial search; 993 were excluded due to unrelated titles. Forty-eight studies were considered for a full-text review, and 29 were excluded based on exclusion criteria. Finally, 19 studies [9-27] were included in the systematic review, and 16 were included in the meta-analysis [9-14, 16, 17, 19, 21-27] [Figure 1].

### Study Characteristics

Studies considered for the systematic review were cross-sectional studies using online self-administered or telephone-administered questionnaires. Among the 19 studies included for this review, six were from China [14, 15, 18, 19, 22, 23], two were from Iran [11, 16], two from US [13, 27], one from (US and UK) [20], two from multiple countries [9, 10], with one each of the following from Vietnam [12], India [17], Bangladesh [21], Turkey [24], Jordan [25] and Malaysia [26]. The sample sizes ranged from 85 [11] to 5974 [20]. The primary target population across studies were general population (n=9) [13, 14, 17, 18, 20-23, 26], healthcare workers [HCWs] (n=8) [9-12, 15, 19, 24, 25], medical students (n=1) [16] and people with chronic disease (n=1) [27]. Additional details are reported in Table 1.

**Table 1.**
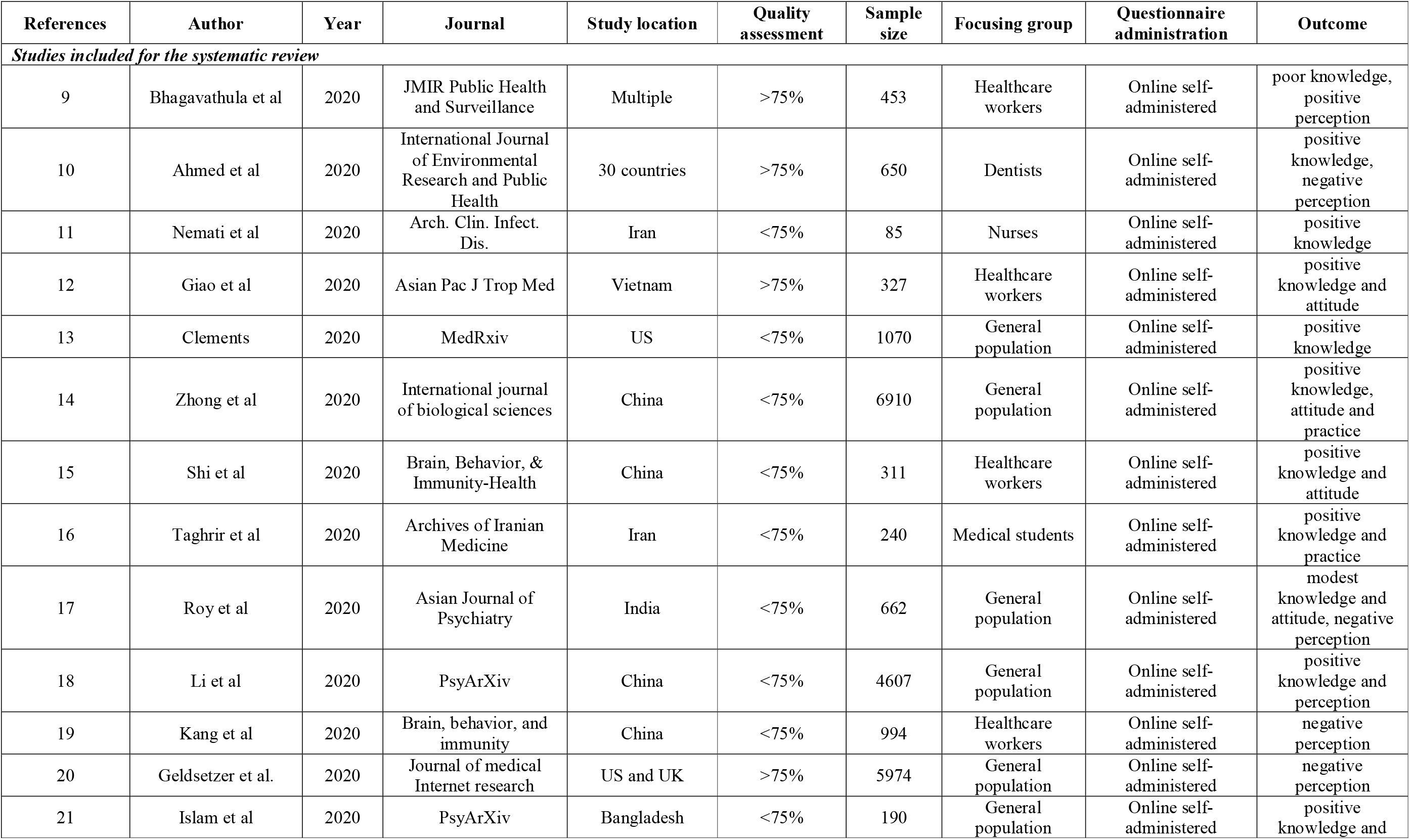
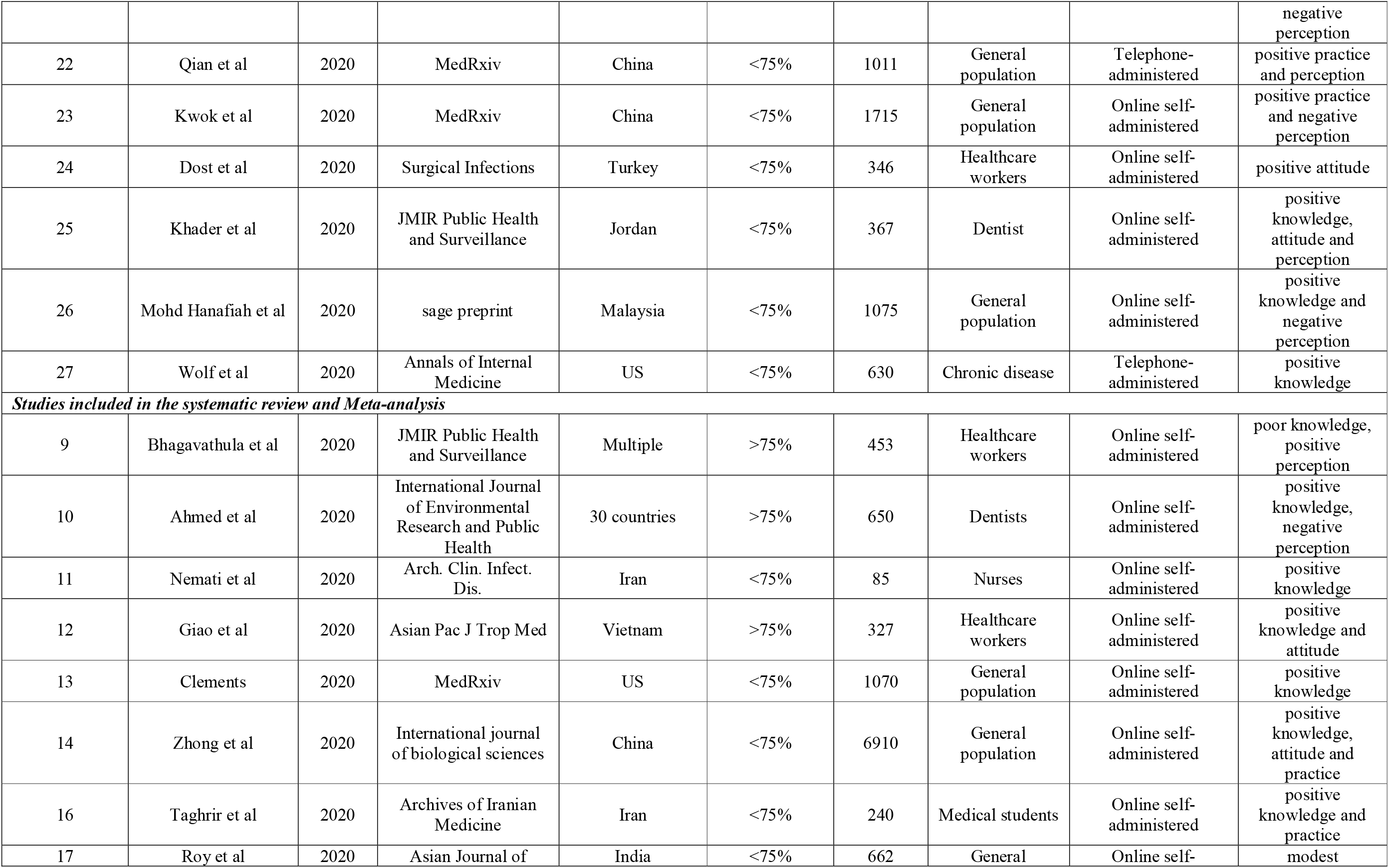
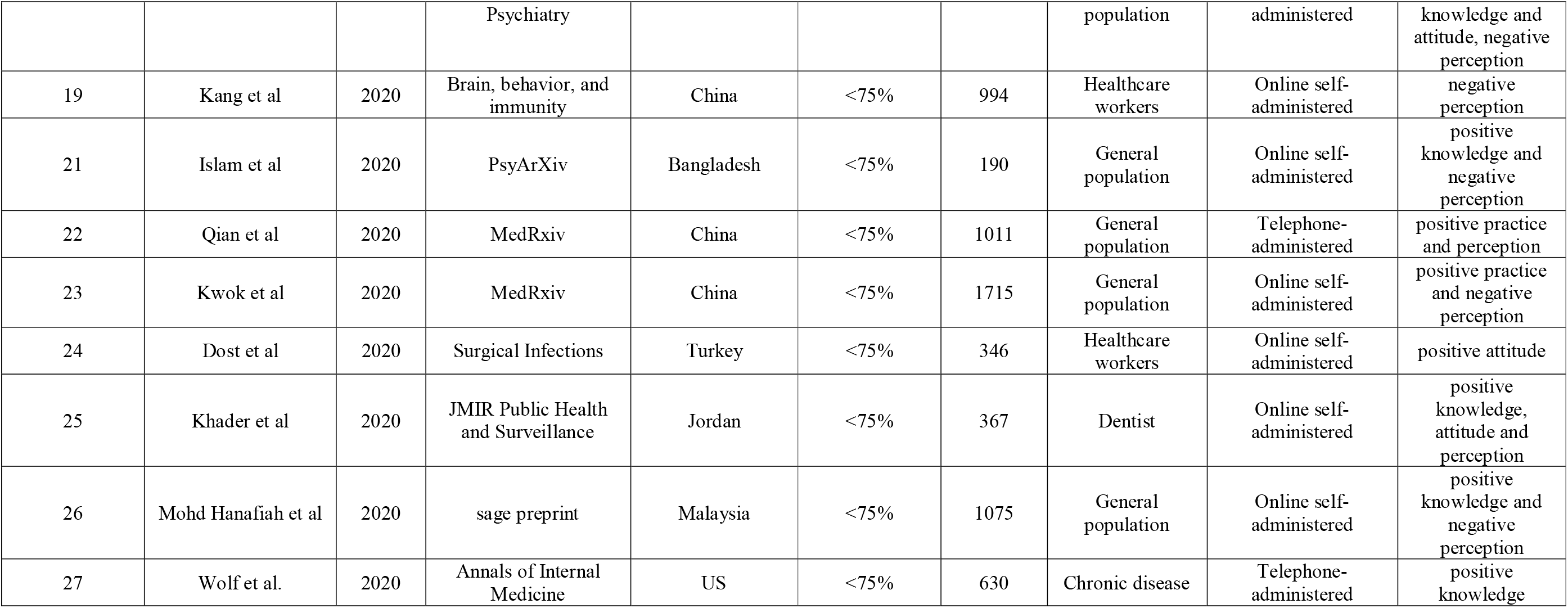
Characteristics of the cross-sectional studies included in Systematic review and Meta-analysis (N=27,617)

### Knowledge about COVID-19

Knowledge about COVID-19 was assessed with two statements and the results are presented in Figures 2 and 3. Nine studies reported the knowledge of COVID-19 symptoms [9, 12-14, 16, 17, 25-27], overall, 79% (95% CI: 69-89) of the sample correctly identified COVID-19 related symptoms. Seven studies reported the knowledge of COVID-19 transmission [9, 10, 12-14, 16, 25] with an overall 82% (95% CI: 74-90) are aware of its transmission. Knowledge about COVID-19 transmission was high among the general population (91.8%) [13, 14] while knowledge about COVID-19 symptoms was high among HCWs and medical students (82.9%) [9, 12, 16, 25] [Table 2].

**Table 2:** Knowledge, attitude, Perception and Practice about COVID-19 among various study groups

|  | No of studies | Sample size | Estimates | 95% CI | P-value | I<sup>2</sup> |
| --- | --- | --- | --- | --- | --- | --- |
| **HCWs and medical students** |  |  |  |  |  |  |
| General knowledge | 1 | 85 | 56.5 | 45.9-67 | - | - |
| Knowledge about transmission | 5 | 2037 | 77.3 | 59.9-94.7 | <0.001 | 99.4% |
| Symptoms | 4 | 1387 | 82.9 | 71.3-94.5 | <0.001 | 97.6% |
| Attitude about COVID-19 isolation | 1 | 327 | 97.9 | 96.3-99.4 | - | - |
| Avoiding crowded areas | 1 | 240 | 99.6 | 98.8-100.4 | - | - |
| Perception about COVID-19 | 4 | 2464 | 58.9 | 13.2-104.7 | <0.001 | 99.9% |
| Hand hygiene can prevent COVID-19 | 3 | 1040 | 76.5 | 58-95 | <0.001 | 98.5% |
| Practice about COVID-19, wearing mask | - | - | - | - | - | - |
| **General population** |  |  |  |  |  |  |
| General knowledge | 1 | 190 | 73.2 | 66.9-79.5 | - | - |
| Knowledge about transmission | 2 | 7980 | 91.8 | 79.9-103.6 | <0.001 | 99.2% |
| Symptoms | 4 | 9717 | 76.7 | 60.9-93.3 | <0.001 | 99.8% |
| Attitude about COVID-19 isolation | 1 | 662 | 95.9 | 94.4-97.4 | - | - |
| Avoiding crowded areas | 3 | 9695 | 88 | 86.5-89.6 | <0.001 | 99.5% |
| Perception about COVID-19 | 5 | 4653 | 35.1 | 5.5-64.7 | <0.001 | 99.8% |
| Hand hygiene can prevent COVID-19 | 1 | 662 | 97 | 95.7-98.3 | - | - |
| Practice about COVID-19, wearing mask | 4 | 10706 | 73.1 | 60.8-85.3 | <0.001 | 99.9% |
| **Patients with Chronic diseases** |  |  |  |  |  |  |
| Symptoms | 1 | 630 | 71.6 | 68.1-75.1 | - | - |

**Figure 2:**
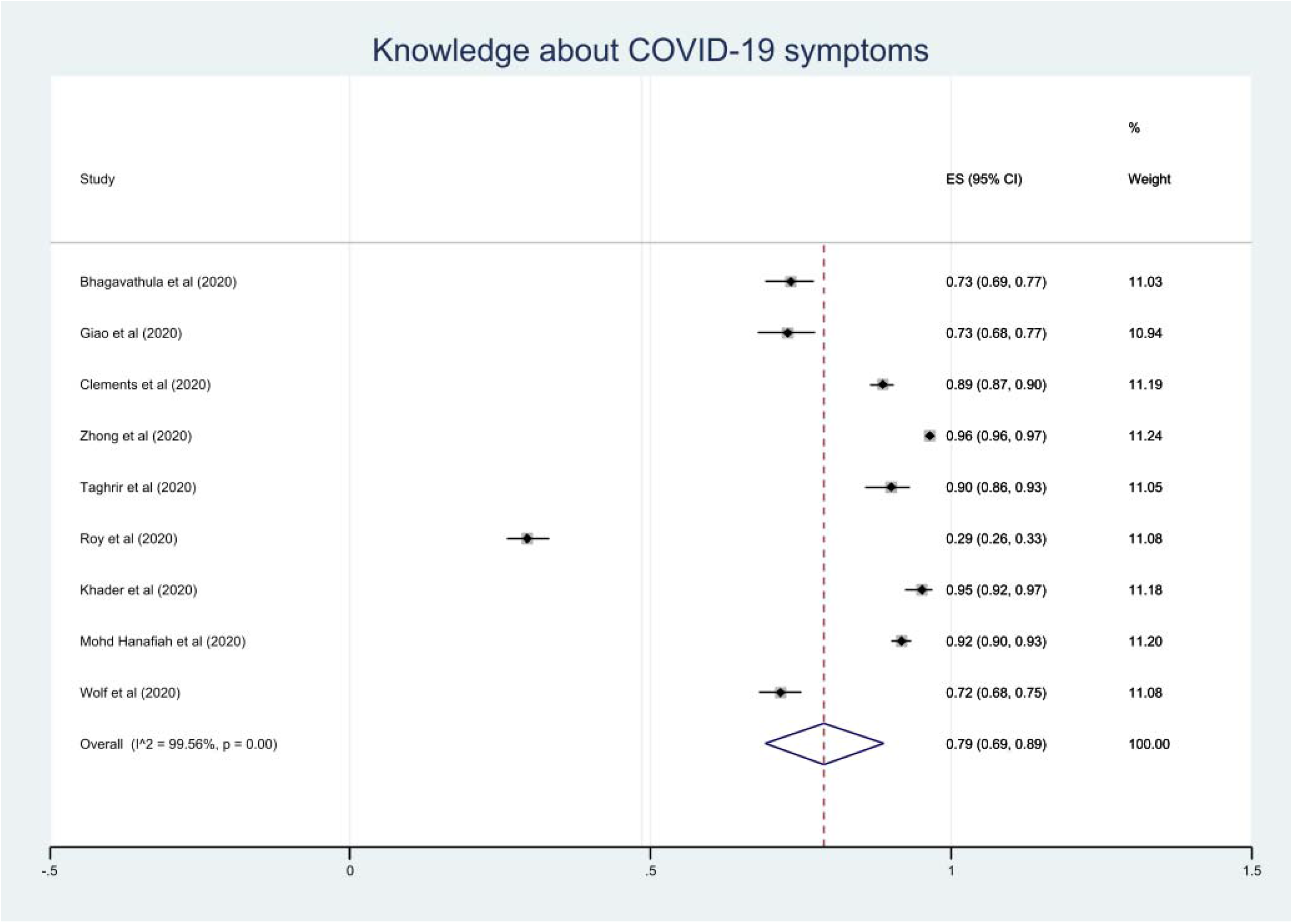
Knowledge about COVID-19-related symptoms.

**Figure 3:**
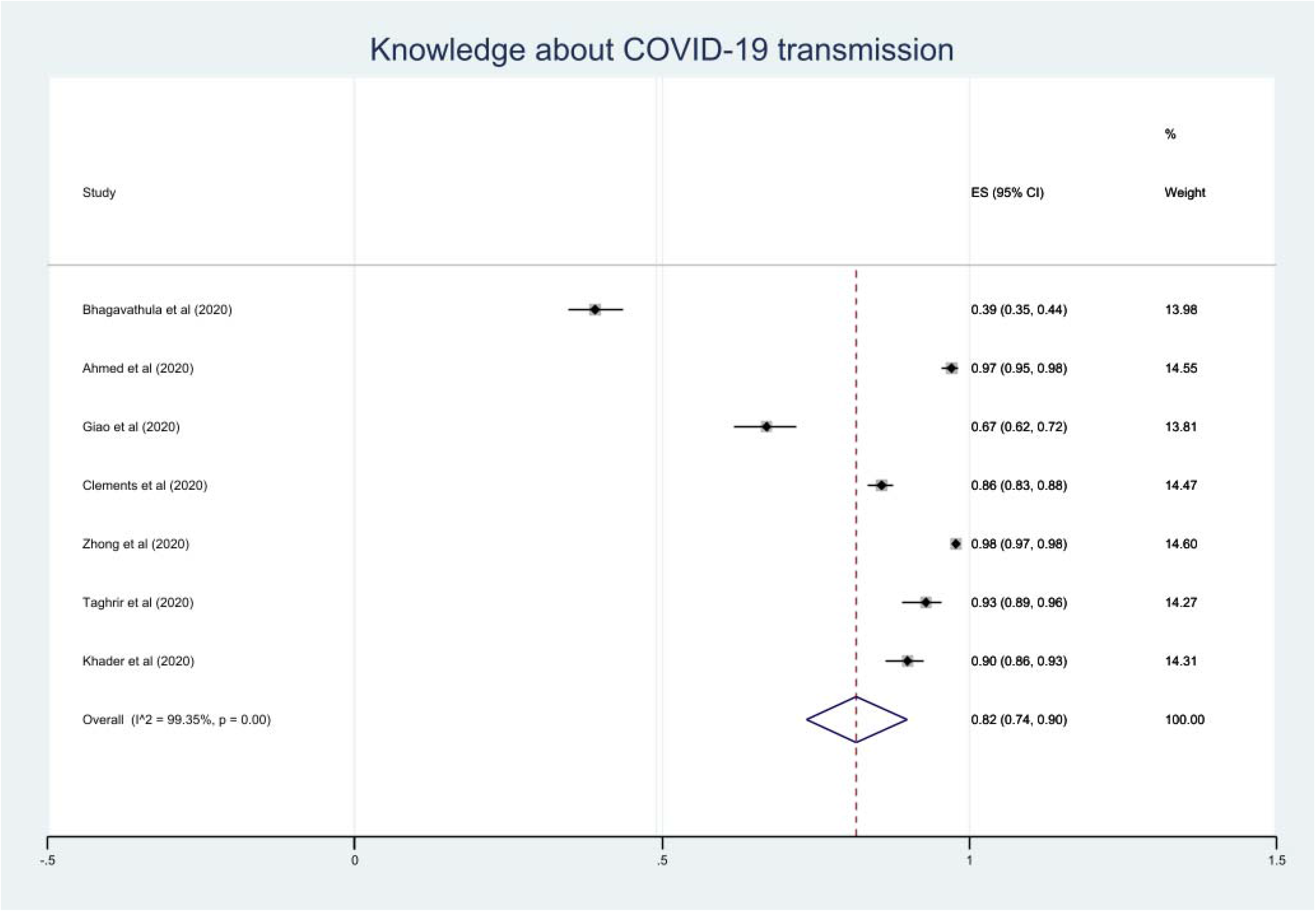
Knowledge about COVID-19 transmission.

### Attitude towards COVID-19

Four studies reported the attitude towards COVID-19 as avoiding crowded areas [13, 14, 16, 23] and 89% (95% CI: 82-95; *p*<0.001) of the participants reported that they avoided crowded places to prevent getting COVID-19 [Figure 4]. Subgroup analysis found that medical students [16], as well as the general population [13, 14, 23], had a positive attitude towards COVID-19 with 99.6% and 88%, respectively. More than 95% of the HCWs (97.9%) and general public (95.9%) opinioned that COVID-19 patients should be isolated (Table 2).

**Figure 4:**
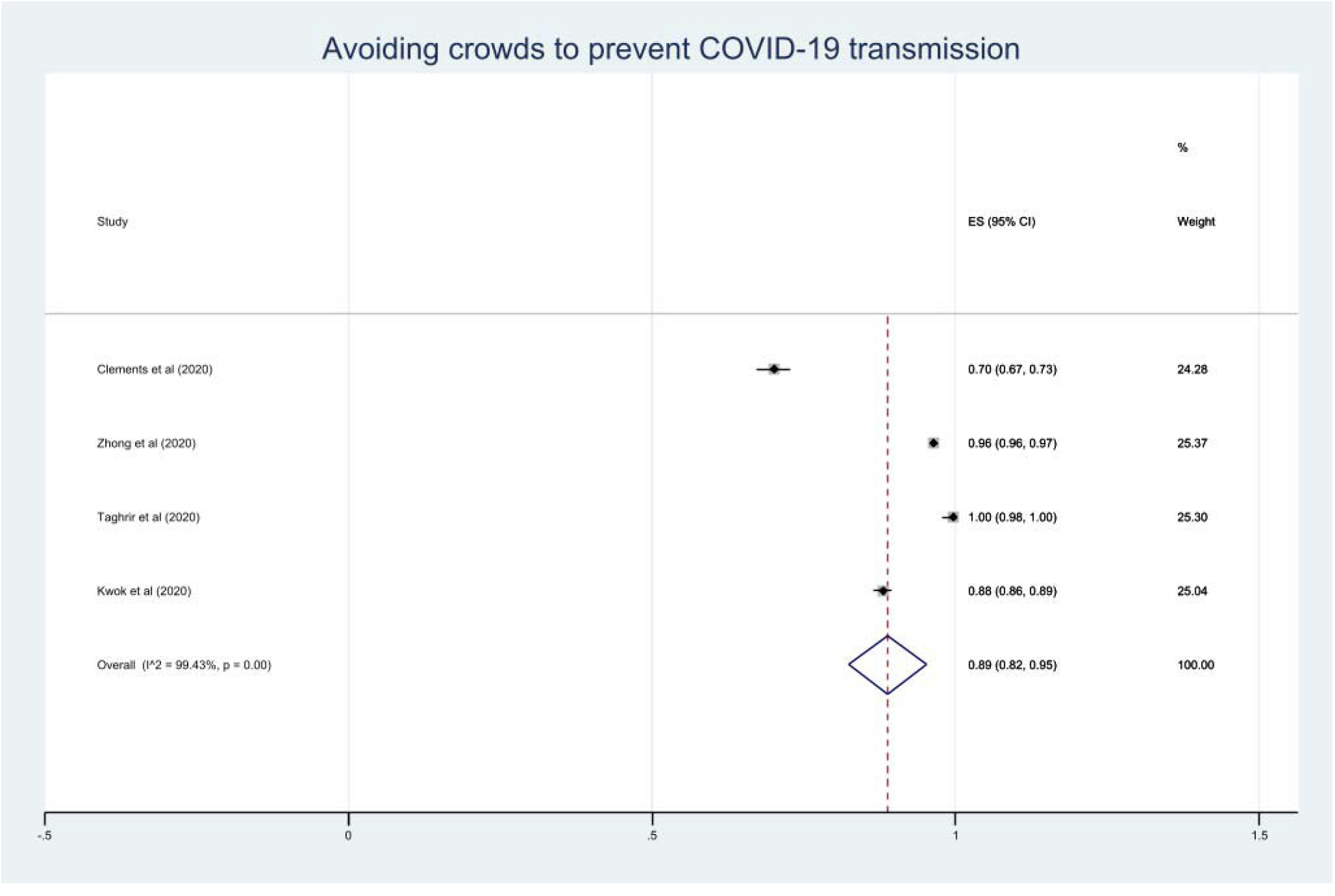
Attitude: Avoid crowded areas to prevent COVID-19 transmission.

### Perception of COVID-19

Nine studies reported perceptions about COVID-19 [9, 10, 17, 19, 21-23, 25, 26]. Overall, only 46% (95% CI: 15-77) of the population studied showed positive perceptions about COVID-19 [Figure 5]. Moreover, these perceptions among HCWs [9, 10, 19, 25] were more positive (58.9%) when compared to the general population (35.1%) [17, 21-23, 26] [Table 2].

**Figure 5:**
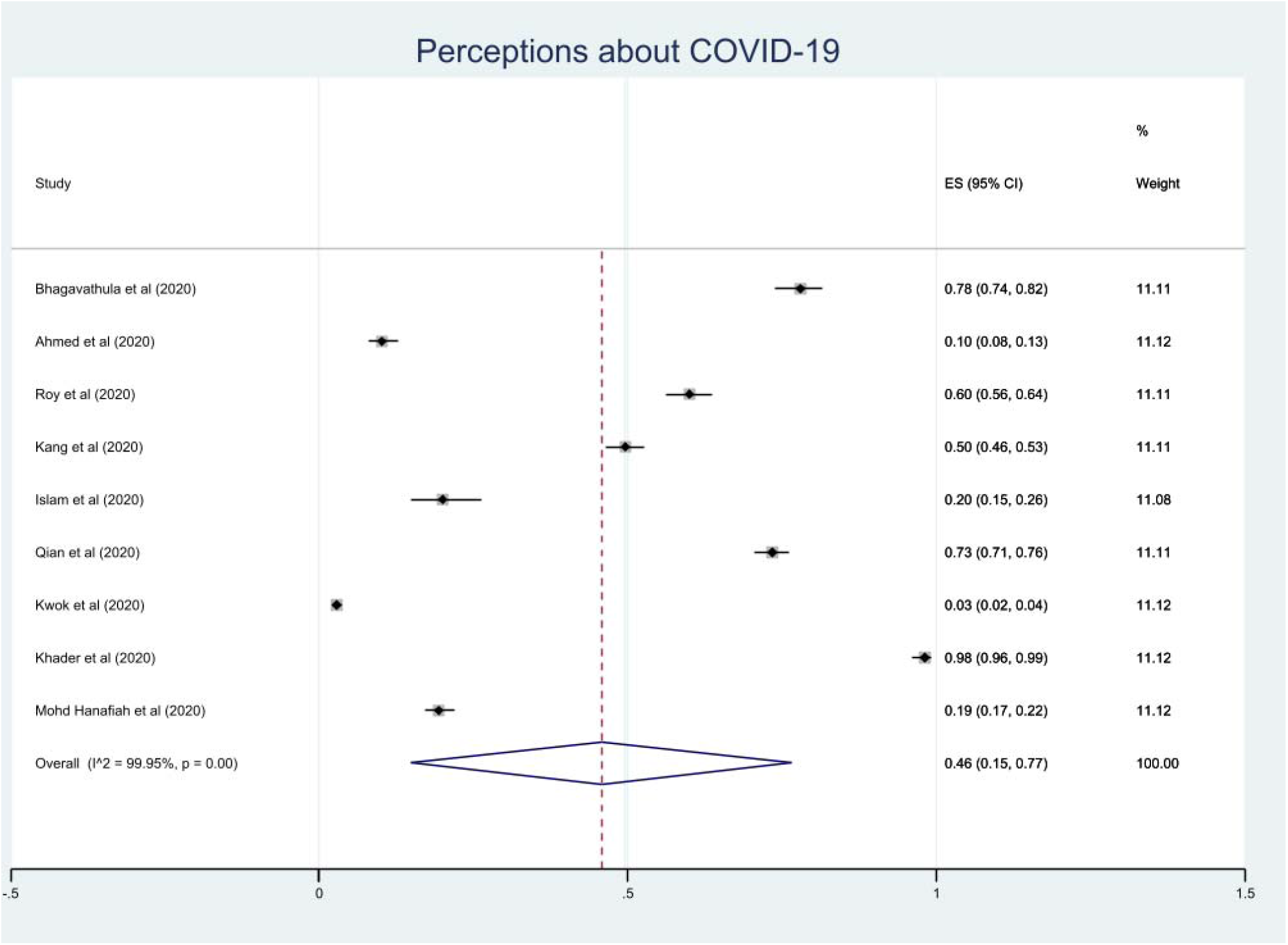
Perceptions about COVID-19.

### Practice on COVID-19 Prevention

Practice towards COVID-19 prevention was assessed using statements in studies related to wearing a face mask and maintaining hand hygiene. Four studies reported practice related to wearing a face mask to prevent COVID-19 transmission [13, 14, 22, 23] with an overall percentage of 73% (95% CI: 61-85) [Figure 6]. For hand hygiene practices, four studies assessed a cumulative sample of 1702 population and 82% (95% CI: 68-95) of the sample agreed that they practice hand hygiene to prevent them from COVID-19 [12, 17, 24, 25] [Figure 7]. Subgroup analysis showed that practices among the general population by wearing face mask [13, 14, 22, 23] and hand hygiene [17] were 73.1% and 97%, respectively. While hand hygiene practice among HCWs [12, 24, 25] and medical students [17] was suboptimal (76.5%) [Table 2]

**Figure 6:**
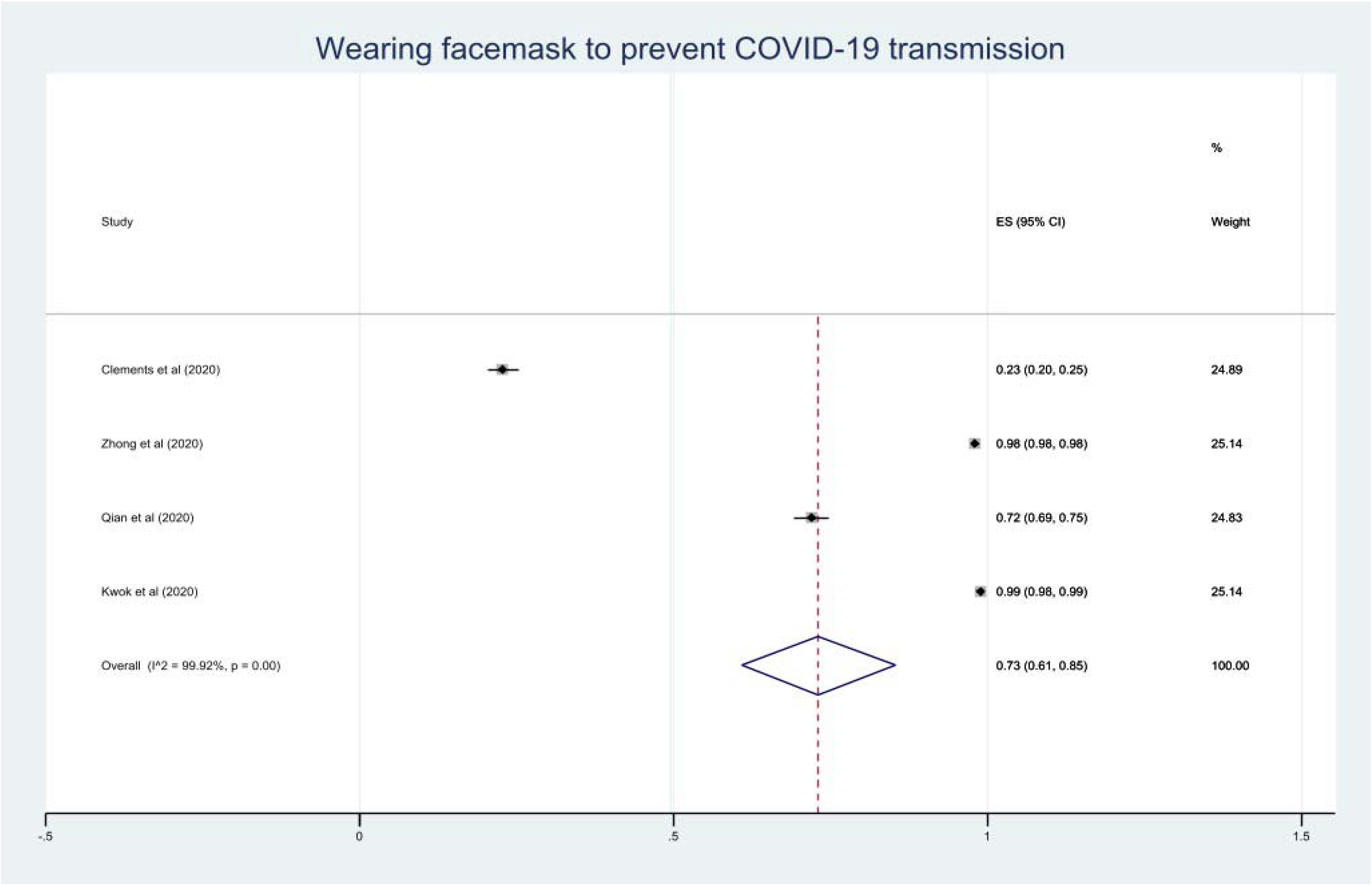
Practice: Wearing face mask to prevent COVID-19 transmission.

**Figure 7:**
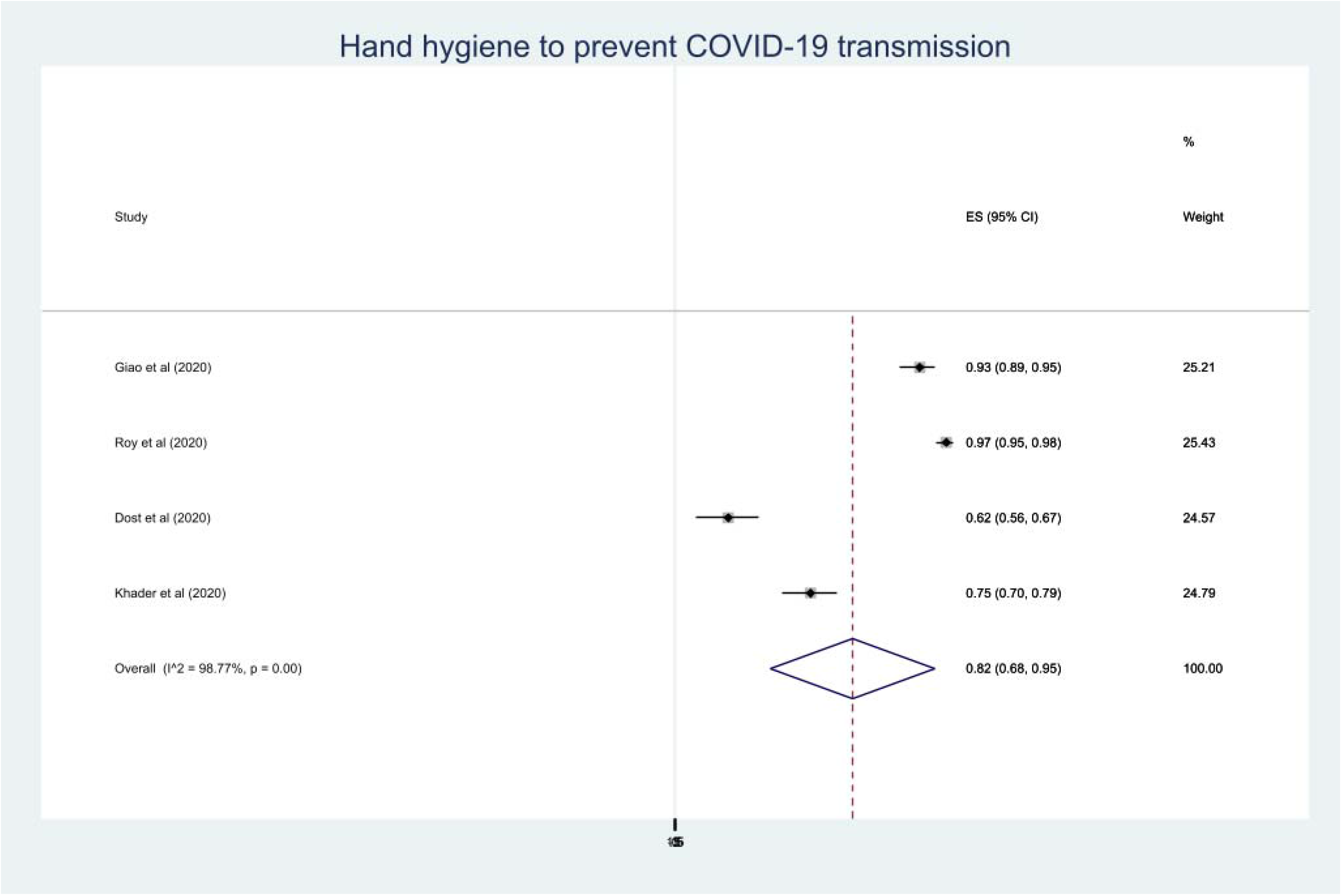
Practice: Hand hygiene practices to prevent COVID-19 transmission.

### Sensitivity analysis

To address the issue of heterogeneity, studies are classified into high (>75%) and low quality (<75%) following the STROBE checklist for methodological quality. No significant differences in the knowledge levels were seen between low- and high-quality studies [Table 3]. Thirteen studies were identified as low-quality at reporting attitude levels [11, 13, 14, 16, 17, 19, 21-27], with only 46.2% of the sample had a positive perception about COVID-19.

**Table 3:**
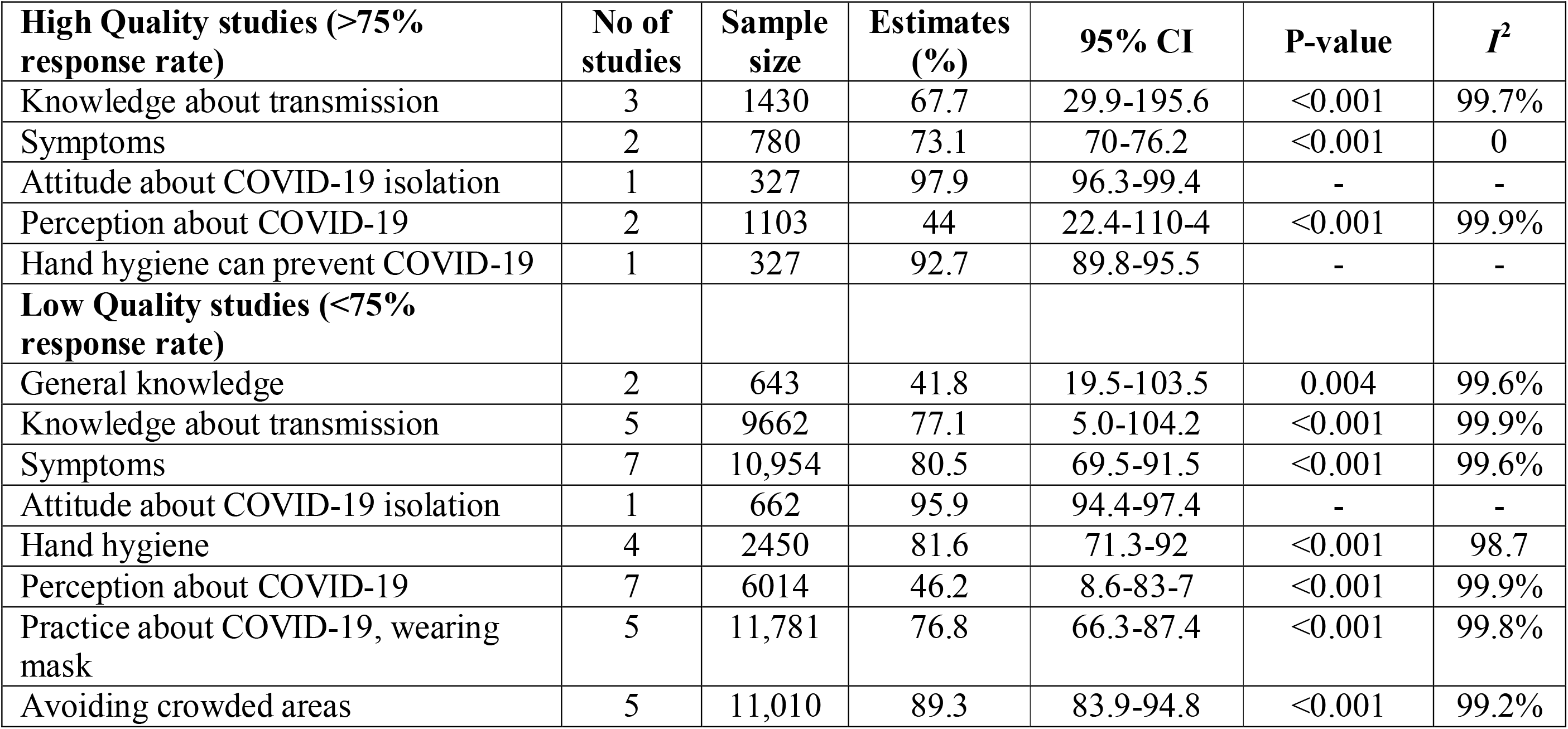
Subgroup analysis: Quality of studies

### Study quality assessment

Study quality was evaluated using the Joanna Briggs Institute’s criteria [Table 4]. A set of nine criteria were used to evaluate the quality of studies. Ten studies showed that the sample represented the target population [9-12, 15, 16, 19, 24, 25, 27] and five studies showed that the participants have been recruited appropriately [9, 12, 18, 22, 25]. Two studies calculated the sample size [16, 24], and 17 studies described their study settings [9-16, 18-20, 22-27]. The majority of the studies conducted the data analysis sufficiently [9-19, 22-27], however, standard criteria were not used to assess the KAP about COVID-19 [9-27]. Some studies measured precisely on HCWs and medical students [9, 10, 16, 19], used appropriate statistical analysis [9, 12-16, 18, 19, 22-24, 26], but none identified major confounders and subgroups [9-27].

**Table 4:** Study quality assessment of included studies that evaluated the knowledge, attitude, perception, and practice towards COVID-19

| Study | Was the sample representative of the target population? | Were study participants recruited in an appropriate way? | Was the sample size adequate? | Were the study subjects and the setting described in detail? | Was the data analysis conducted with sufficient coverage of the identified sample? | Were objective, standard criteria used for the measurement of the condition? | Was the condition measured reliably? | Was there appropriate statistical analysis? | Are all the important confounding factors/subgroups/differences identified and accounted for? |
| --- | --- | --- | --- | --- | --- | --- | --- | --- | --- |
| **Bhagavathula et al** | Yes | Yes | Not clear | Yes | Yes | No | Yes | Yes | No |
| **Ahmed et al** | Yes | No | Not clear | Yes | Yes | No | Yes | No | No |
| **Nemati et al** | Yes | No | Not clear | Yes | Yes | No | No | No | No |
| **Giao et al** | Yes | Yes | Not clear | Yes | Yes | No | No | Yes | No |
| **Clements** | No | No | Not clear | Yes | Yes | No | No | Yes | No |
| **Zhong et al** | No | No | Not clear | Yes | Yes | No | No | Yes | No |
| **Shi et al** | Yes | No | Not clear | Yes | Yes | No | No | Yes | No |
| **Taghrir et al** | Yes | No | Yes | Yes | Yes | No | Yes | Yes | No |
| **Roy et al** | No | No | Not clear | No | Yes | No | No | No | No |
| **Li et al** | No | Yes | Not clear | Yes | Yes | No | No | Yes | No |
| **Kang et al** | Yes | No | Not clear | Yes | Yes | No | Yes | Yes | No |
| **Geldsetzer et al** | No | No | Not clear | Yes | No | No | No | No | No |
| **Islam et al** | No | No | Not clear | No | No | No | No | No | No |
| **Qian et al** | No | Yes | Not clear | Yes | Yes | No | No | Yes | No |
| **Kwok et al** | No | No | Not clear | Yes | Yes | No | No | Yes | No |
| **Dost et al** | Yes | No | Yes | Yes | Yes | No | No | Yes | No |
| **Khader et al** | Yes | Yes | Not clear | Yes | Yes | No | No | No | No |
| **Mohd Hanafiah et al** | No | No | Not clear | Yes | Yes | No | No | Yes | No |
| **Wolf et al** | Yes | No | Not clear | Yes | Yes | No | No | No | No |

Abbreviation: COVID-19; Coronavirus disease 2019

### Publication Bias

Publication bias was highlighted in all the 16 studies and was confirmed by asymmetric funnel plots (Supplementary file). Further investigating the extent of publication bias across the statements, the Egger test identified a considerable proportion of bias identified in the knowledge statements related to COVID-19 transmission and COVID-19 symptoms (*p*<0.05). However, the Begg test did not identify any publication bias across the statements (*p*>0.05) [Table 5].

**Table 5:** Risk of bias

| **COVID-19-related** | **Egger test** |  | **Begg test** |  |
| --- | --- | --- | --- | --- |
|  | *t-value* | *P-value* | *z-value* | *P-value* |
| Transmission | -2.81 | **0.03** | 1.80 | 0.072 |
| Symptoms | -2.73 | **0.029** | 1.15 | 0.251 |
| Perceptions | 1.21 | 0.266 | 0.83 | 0.73 |
| Avoiding crowds | -1.68 | 0.234 | 1.02 | 0.308 |
| Hand hygiene | -3.72 | 0.061 | 1.70 | 0.089 |
| Wearing face masks | -2.27 | 0.151 | 0.34 | 0.734 |

## Discussion

In this first and largest review, we assess the nature and extent of KAP towards COVID-19 during the pandemic period from January to May 31, 2020. Two recent reviews focused on understanding the extent of preparedness of medical students towards disaster training programs [32] and efficacy of face mask in preventing respiratory virus [33]. Absent a vaccine or drug therapy to prevent and cure COVID-19 infection, there is irrefutable evidence on prevention of transmission by public health measures such as handwashing, wearing masks, and practicing social distancing. In this review of 19 studies [9-27], results provide robust evidence about KAP gaps in COVID-19 infection. For example, we found that the majority of the sample had good knowledge of COVID-19 clinical symptoms (79%) and its transmission (82%), and the majority (89%) agreed that individuals should avoid going to crowded places to prevent COVID-19 transmission. Furthermore, more than 50% of the cumulative sample had negative perceptions about COVID-19. Overall, nearly three-fourth (73%) of the sample indicated that COVID-19 prevention practices such as wearing face masks and maintaining hand hygiene should be prioritized to prevent its transmission.

People’s knowledge, attitude, perceptions, and practices about COVID-19 are important predictors of whether they engage in disease-specific preventive behaviors. Evidence from prior studies indicates that higher knowledge about infectious diseases is positively associated with increased engagement in appropriate protective behavior during an outbreak [34-37]. However, widespread endorsement of medical conspiracy beliefs to counter scientific evidence can inhibit preventive behavior [38-40]. In several instances, knowledge can also influence perceptions due to past experiences and beliefs [40-42]. The findings of this study suggest a significant gap between the amount of knowledge, attitude, and prevention practices related to COVID-19 infection. This is a critical finding as the perceptions of COVID-19 could be important predictors of whether people engage in disease-specific preventive behaviors or not. Furthermore, the knowledge and perceptions share some of the same psychological motivations such as perceived risk and self-efficacy to control COVID-19. Although we did not investigate the underlying reasons for variations in risk perceptions, additional research is warranted based on psychological theories and health behavior change models to explore and explain the connections between KAP and COVID-19 preventive behaviors such as mask-wearing.

The findings of this review specifically point to a widespread lack of practice of mask-wearing to prevent COVID-19 among study subjects. Wearing a face mask as appropriate is important to prevent transmission of COVID-19 infections and can act as a physical barrier to the spread of droplets. While mask-wearing was not recommended initially by health agencies such as the World health organization [43] and US Center for Disease Control [44] as a key strategy to reduce transmission of COVID-19 in the community, continuing accumulation of evidence now suggests that mask-wearing is one of the key preventive behaviors to halt the community-based spread of the infection [45-47].

A major disconcerting finding of this review pertains to HCWs and medical students. One would assume that these groups of individuals would have a better outlook and behaviors regarding COVID-19 given their field of training and expertise. However, our subgroup analysis found that the level of knowledge on the disease (56.5%) and perceptions (58.9%) about COVID-19 among HCWs and medical students are suboptimal. Such a deficit during an ongoing pandemic may result in delayed recognition and handling of potential COVID-19 patients. Furthermore, if these perceptions are truly representative, this could have an impact on patient care and also on the dynamics of the COVID-19 outbreak. There is an urgent need to develop evidence-based educational interventions to improve HCWs level of knowledge and to alter their KAP.

## Limitations

The results of this review must be considered in light of several potential limitations. The meta-analysis shows high heterogeneity with a lack of homogeneity of the responses of participants among various studies. This could be due to sociodemographic and socio-cultural variations among different study subjects and differences in questionnaire content and administration. These issues may have led to selection bias. Moreover, we used a set of questions to quantify the level of KAP about COVID-19 and published until the end of May 2020 are included. Thus, our analysis may have missed additional findings from the literature. Furthermore, our analysis used secondary data in the published literature that some of the responses may overestimate or underestimate while providing results that might lead to reporting or recall bias. The Quality assessment of each included study allowed us to evaluate the presence of potential bias and confounding. Also, funnel plots revealed asymmetry across all outcomes. Thus, caution should be taken while interpreting the results of this review. Despite this, no publication bias was identified by the Begg test. Lastly, this review has the largest sample size, with the latest findings, and covers the widest geography to date on the topics under investigation.

## Conclusion

Our results identified a high majority of the sample exhibited negative perceptions about COVID-19 and preventive practice of wearing face masks and maintaining hand hygiene are suboptimal. Most of the HCWs and medical students have poor knowledge and two-third (65%) of the general public have a negative attitude towards COVID-19. Reinforcing evidence-based educational interventions among HCWs and reinforcing prevention guidelines to improve their understanding and thus can alter the KAP in the general population during the COVID-19 era.

## Data Availability

All the data is available within the manuscript

## Acknowledgment

We thank PROSPERO for reviewing the protocol and providing their support in fulfilling the research work.

## Authors’ contributions

All the authors equally contributed in literature review, data collection, analysis and writing the manuscript. All authors have read and approved the final manuscript.

## Available data and materials

All materials are attached as supplementary materials, and information related to the study is in the manuscript.

